# LLM Representations Rival Traditional Predictors and Enhance Multimodal Prediction of APOE4 and Aβ-Burden

**DOI:** 10.64898/2026.11.09.26367620

**Authors:** Maya Zadok, Nitay Calderon, Yoav Wald, Netaniel Rein, Ithamar Ganmore, Orit Lesman Segev, Ganit Almog, Tamar Shamir, Rotem Ackler, Shaked Y. Leviner, Gal Noimark, Gil Levy, Kimberly Diggle Mueller, Ramit Ravona Springer, Michal Schnaider Beeri, Roi Reichart

## Abstract

Spontaneous language offers a scalable, non-invasive approach for identifying biological indicators of preclinical Alzheimer’s disease (AD). We examined prediction of APOE4 carrier status and cerebral amyloid-β (Aβ) burden using Cookie Theft picture-descriptions from two cohorts, including Hebrew- and English-speakers (IRAP, N=134; and WRAP, N=1,541). Aβ burden was classified across lowest-quartile, median, and highest-quartile cutoffs, capturing accumulation below conventional amyloid-positivity thresholds. We compared embeddings from pretrained large language models (LLMs) to standard cognitive test scores, reflecting current clinical assessment, and theory-driven linguistic features, reflecting traditional language analysis. Embedding-based models achieve the highest F1 among constituent models in five of eight tasks for APOE4 status and global Aβ. High prediction disagreement suggests complementary signals between approaches. An any-positive ensemble significantly outperforms every constituent model and a prior-informed random baseline across all eight tasks, and is also significantly superior to all constituent models in 21 of 24 regional Aβ tasks. These findings highlight the complementarity of LLM-based, theory-driven, and cognitive measures for predicting AD-related genetic risk and pathology, and supports development of early screening approaches.

## 1 Introduction

Alzheimer’s disease (AD) is a neurodegenerative disorder and the most common cause of dementia. Its pathological processes can begin many years before clinically apparent symptoms, creating an important challenge for early detection and intervention. Early identification is particularly important because available treatments and preventive measures are most effective before substantial cognitive deterioration has occurred^1–3^. However, current diagnostic pathways are often initiated only after symptoms become noticeable, when the underlying pathology may already have been developing for decades^4^. This motivates the search for accessible, non-invasive methods that can identify signals associated with AD risk and pathology at early stages.

Language is a particularly promising source of information for this purpose. Language production involves multiple cognitive processes, and changes in linguistic abilities have long been recognized as characteristic of AD progression. Indeed, language-related tasks are central components of standard neuropsychological assessments, (e.g., verbal fluency, naming, and verbal memory tests^5–7^). Beyond these structured assessments, spontaneous speech provides an opportunity to examine how individuals produce language in a more naturalistic setting. One widely used approach is the Cookie Theft picture-description task^8^, in which participants describe a standardized visual scene. The simplicity of collecting such speech samples, together with advances in Artificial Intelligence (AI), has motivated substantial research into language-based indicators of cognitive impairment and dementia.

Earlier studies investigated linguistic changes through manual annotation and relatively simple quantitative measures^9–11^. More recent work has employed Natural Language Processing (NLP) and machine learning to extract linguistic information automatically and predict cognitive status from speech. These approaches include models based on predefined, theory-driven linguistic features, such as lexical diversity, syntactic complexity, and word frequency^12–17^, as well as models that learn representations of language directly from data^18–23^. The emergence of Large Language Models (LLMs) has substantially expanded the possibilities for the latter approach. Trained on very large text corpora to predict and generate language and perform a broad range of linguistic tasks, LLMs develop internal representations that capture rich aspects of linguistic meaning and context. These representations offer a promising means of identifying subtle language patterns that may be difficult to specify in advance. LLMs can also support automatic extraction of complex linguistic features that typically require human judgment, reducing the need for extensive manual annotation.

Despite this progress, an important scientific and clinical challenge remains. Much of the research on language-based AD prediction studies have focused on distinguishing individuals with dementia or Mild Cognitive Impairment (MCI) from cognitively healthy controls^24^. Although these studies have established that language contains useful information about cognitive impairment, they leave open a more fundamental question: Can language provide information about AD-related biological risk and pathology before clear clinical symptoms emerge? Addressing this question requires moving beyond the prediction of cognitive status toward biological indicators associated with the earliest stages of the disease.

In this work, we focus on two such indicators: apolipoprotein E ε4 (APOE4) carrier status and cerebral amyloid-β (Aβ) burden. APOE4 is the strongest common genetic risk factor for late-onset AD, with carriers facing an elevated risk of developing the disease^3,25^. Aβ accumulation, in turn, is a central feature of AD neuropathology and can begin long before the onset of overt cognitive impairment^26^. These indicators capture different aspects of AD: APOE4 reflects inherited susceptibility, whereas Aβ burden provides a measure of underlying pathological processes. Investigating their relationships with language therefore offers an opportunity to examine whether linguistic behavior contains information associated with both biological vulnerability and disease pathology at early stages.

Our central hypothesis is that different representations of language and cognition capture distinct, potentially complementary signals associated with these biological indicators. In particular, we consider three sources of information: traditional cognitive assessment scores, which serve as our primary baseline; theory-driven (top-down) linguistic features extracted from spontaneous speech, which reflect the traditional approach to language analysis and serve as a second baseline; and language representations derived from pretrained LLMs. These approaches characterize individuals from different perspectives. Cognitive assessments measure performance on established neuropsychological tasks; predefined linguistic features capture specific aspects of language production and content; and LLM representations provide a broader, data-driven characterization of linguistic information. Although all three may be informative about AD-related risk and pathology, there is no reason to assume that they capture the same information or succeed and fail on the same individuals.

We investigate this hypothesis using data from two longitudinal cohorts: the Wisconsin Registry for Alzheimer’s Prevention (WRAP)^27^ and the Israel Registry for Alzheimer’s Prevention (IRAP)^28^. Both cohorts include individuals at elevated risk for AD and provide cognitive assessments alongside genetic and neuroimaging information. We analyze spontaneous speech elicited through the Cookie Theft task, using English-language data from WRAP and Hebrew-language data collected in IRAP specifically for this study. The inclusion of these two cohorts allows us to examine the proposed approach across different languages and populations, while also extending computational language analysis to a Hebrew-speaking population that remains underrepresented in the AD literature^29–31^.

We develop separate predictive models based on each of the three sources of information using Hierarchical Multi-Domain Regression (HMDR), a linear modeling framework that estimates shared and domain-specific effects. Importantly, rather than considering amyloid pathology only through a conventional positive-versus-negative distinction, we also examine multiple levels of amyloid accumulation, including levels below established clinical positivity thresholds. This allows us to investigate whether the available linguistic and cognitive signals are informative about variation in Aβ burden during earlier stages of the pathological process. We apply this approach to APOE4 carrier status and to global amyloid measures, and further extend the amyloid analysis to regional burden in the cingulate, frontal, parietal and temporal cortices to assess whether predictive patterns differ across brain regions.

Our results reveal that the three models achieve broadly comparable predictive performance, but their predictions differ substantially at the individual level. Across tasks and cohorts, observations correctly classified by one model are frequently misclassified by another, and no single representation consistently dominates. These findings provide evidence that the three sources of information capture complementary signals associated with APOE4 status and Aβ burden. Motivated by this complementarity, we investigate whether the different signals can mutually enhance prediction. We combine the three models using a simple ensemble that identifies a positive case whenever at least one constituent model predicts a positive outcome. This ensemble is the best-performing model, significantly improving F1 relative to each individual model for both APOE4 and global amyloid in both cohorts, demonstrating the predictive value of integrating information from language and cognitive assessments. The ensemble benefit also extends to regional amyloid outcomes, with significant advantages over all three constituent models in 21 of 24 tasks defined across the four cortical regions.

Together, these findings suggest that spontaneous language contains information associated with AD-related genetic risk and amyloid pathology that is not fully captured by standard cognitive assessments. They also demonstrate that theory-driven linguistic features and LLM-based representations provide complementary information, despite being derived from the same speech samples. More broadly, our work provides evidence for the potential of combining modern language-based AI methods with established cognitive measures to improve the identification of biological indicators associated with early AD. This offers a foundation for developing accessible, low-burden screening approaches and for enriching existing clinical assessments with computational analysis of spontaneous speech.

## 2 Results

### 2.1 Sample Characteristics

Table 1 summarizes participant characteristics at the time of the included recordings. The cohorts did not differ significantly in age, sex distribution, or the scan-speech gap (age: t(160.2) = -0.60, p=0.547; sex: χ²(1) = 2.07, p=0.150; scan-speech gap: t(69.6) = 1.36, p=0.178). Mean years of education were slightly higher in WRAP than in IRAP (16.6 vs. 16.0 years, *t(167.6) = -2.68, p*=0.008). WRAP also had a higher proportion of native speakers (97.6% vs. 79.9%, *χ²(1) = 105.33, p*<0.001) and higher mean Mini-Mental State Examination (MMSE) scores (29.4 vs. 28.6, *t(143.1) = -6.05, p*<0.001). Despite the higher mean MMSE, cognitive impairment was more frequent in IRAP than in WRAP (10.4% vs. 3.5%, *χ²(1) = 15.26, p*<0.001; see <u>Methods</u>). In task-specific comparisons (Supplementary Tables 1 and 2), no significant differences in cognitive impairment were observed between groups in IRAP, whereas in WRAP, the positive groups in the PiB median and Q4 tasks had significantly higher proportions of cognitively impaired participants than the corresponding negative groups.

**Table 1:** Participant Characteristics for Included Recordings Continuous variables (age, education years, MMSE, scan-speech gap) are presented as mean (SD) and were compared using two-sided Welch’s t-test. Categorical variables (sex, native speakers, cognitive status) are presented as n (%) and were compared using Pearson’s chi-square test. Statistically significant results (p<0.05) are shown in bold.

| Variable | IRAP | WRAP | Statistic (df) | p |
|---|---|---|---|---|
| N | 134 | 1541 |  |  |
| Age, mean (SD) | 63.9 (6.9) | 64.3 (7.3) | t(160.2) = -0.60 | 0.547 |
| Female, n (%) | 84 (62.7%) | 1059 (68.7%) | χ²(1) = 2.07 | 0.150 |
| **Education, years, mean (SD)** | **16.0 (2.3)** | **16.6 (2.8)** | **t(167.6) = -2.68** | **0.008** |
| **Native speakers, n (%)** | **107 (79.9%)** | **1500 (97.6%)** | **χ²(1) = 105.33** | **<0.001** |
| NA, n | 0 | 4 |  |  |
| **MMSE, mean (SD)** | **28.6 (1.5)** | **29.4 (1.0)** | **t(143.1) = -6.05** | **<0.001** |
| **Cognitively impaired, n (%)** | **14 (10.4%)** | **54 (3.5%)** | **χ²(1) = 15.26** | **<0.001** |
| Scan-speech gap, years, mean (SD) | 3.8 (1.7) | 3.5 (2.3) | t(69.6) = 1.36 | 0.178 |
| NA, n | 75 | 359 |  |  |

**Table 2:** F1 Scores of Constituent Models.

| **Cohort** | **Task** | **Cog**<br>**[95% CI]** | **Emb**<br>**[95% CI]** | **Top-down**<br>**[95% CI]** |
|---|---|---|---|---|
| **IRAP** | **APOE4** | 0.274<sup>***</sup><br>[0.26, 0.28] | 0.337<br>[0.33, 0.35] | **0.338**<br>**[0.33, 0.35]** |
|  | **SUVR Q1**<br>**(≤0.910)** | **0.334**<br>**[0.32, 0.35]** | 0.285<sup>***</sup><br>[0.27, 0.30] | 0.294<sup>***</sup><br>[0.28, 0.31] |
|  | **SUVR Median**<br>**(>0.941)** | 0.513<sup>***</sup><br>[0.50, 0.52] | **0.536**<br>**[0.53, 0.55]** | 0.494<sup>***</sup><br>[0.48, 0.50] |
|  | **SUVR Q4**<br>**(>0.980)** | 0.205<sup>***</sup><br>[0.19, 0.22] | **0.259**<br>**[0.25, 0.27]** | 0.207<sup>***</sup><br>[0.19, 0.22] |
| **WRAP** | **APOE4** | 0.260<sup>***</sup><br>[0.26, 0.26] | **0.294**<br>**[0.29, 0.30]** | 0.263<sup>***</sup><br>[0.26, 0.27] |
|  | **PiB Q1**<br>**(≤1.046)** | 0.180<sup>***</sup><br>[0.18, 0.18] | **0.281**<br>**[0.28, 0.28]** | 0.260<sup>***</sup><br>[0.26, 0.26] |
|  | **PiB Median**<br>**(>1.092)** | 0.460<sup>***</sup><br>[0.46, 0.46] | **0.475**<br>**[0.47, 0.48]** | 0.450<sup>***</sup><br>[0.45, 0.45] |
|  | **PiB Q4**<br>**(>1.236)** | **0.290**<br>**[0.29, 0.29]** | 0.214<sup>***</sup><br>[0.21, 0.22] | 0.157<sup>***</sup><br>[0.15, 0.16] |

Mean F1 scores across 1,000 MCCV repetitions with 95% confidence intervals. Bold indicates the best-performing approach within each row. Asterisks denote statistically significant differences from the best-performing approach for the corresponding task, based on two-sided paired t-tests with Benjamini-Hochberg (BH) False Discovery Rate (FDR) correction applied to the two comparisons within each task (\* q<0.05, \*\* q<0.01, \*\*\* q<0.001). Test statistics and exact q values are reported in Supplementary Table 6.

### 2.2 Embedding-Based Models Lead Across Tasks, Yet No Single Constituent Model Consistently Dominates

For the eight primary APOE4 and global amyloid tasks, classification performance was evaluated using 1,000 iterations of stratified Monte Carlo Cross-Validation (MCCV), each involving an 80/20 train-test split (see Section 4.3.3). Global Aβ burden was quantified using the Standardized Uptake Value Ratio (SUVR) in IRAP and the Pittsburgh Compound B (PiB) Index in WRAP, and was classified using cohort-specific thresholds for the lowest quartile (Q1), the median, and the highest quartile (Q4) (see <u>Methods 4.2.2</u>). The three constituent models showed broadly comparable classification performance, although their relative ranking varied across prediction tasks (Table 2). The embedding-based model achieved the highest reported F1 score among constituent models in five of the eight tasks. The cognitive model performed best in IRAP SUVR Q1 and WRAP PiB Q4, whereas the top-down linguistic model performed best in IRAP APOE4, although its F1 score was nearly identical to that of the embedding-based model (0.338 vs. 0.337) and the difference was not statistically significant. Thus, while embedding-based representations generally performed best, no single source of features consistently outperformed the others across both cohorts and all tasks.

Cognitive features were selected based on previous literature specific to each cohort, resulting in different sets of cognitive measures for IRAP and WRAP (see Supplementary Methods 1.1). However, all cognitive tests included in the IRAP battery were also available in WRAP, enabling an additional harmonized analysis using a common set of cognitive measures across both cohorts (Supplementary Methods 1.2 and Supplementary Tables 3-5). The harmonized analysis yielded a largely unchanged pattern of results, with no consistently superior constituent model; the best-performing constituent model changed only for IRAP SUVR Q4, where the cognitive model nominally surpassed the embedding model (0.260 vs. 0.259; not statistically significant). The only other change in ranking was a reversal of the cognitive and top-down models, which were second and third, in WRAP PiB Median (0.450 vs. 0.453).

Mean F1 scores across 1,000 MCCV repetitions with 95% confidence intervals. Bold indicates the best-performing approach within each row. Asterisks denote statistically significant differences from the best-performing approach for the corresponding task, based on two-sided paired t-tests with Benjamini-Hochberg (BH) False Discovery Rate (FDR) correction applied to the two comparisons within each task (* q<0.05, ** q<0.01, *** q<0.001). Test statistics and exact q values are reported in Supplementary Table 6.

### 2.3 Individual-Level Predictions Differ Across Constituent Models

Aggregate performance metrics, such as F1 scores, do not reveal whether models correctly classify or misclassify the same observations. We therefore examined the existing predictions of the three constituent models (cognitive, embedding, and top-down) for each observation across all MCCV test sets. Table 3 presents the proportions of agreement and disagreement among these predictions across all test-set observations. Agreement was defined as cases in which all three models made the same prediction, whether correct or incorrect, whereas disagreement occurred when only one or two models correctly classified an observation. To capture both the overall extent of disagreement and its composition, we distinguished four mutually exclusive categories according to the number of models making correct predictions (none, one, two, or all three).

**Table 3:** Distribution of Correct Predictions by Model Agreement Percentages represent the proportion of correct or incorrect predictions across test-set observations over 1,000 MCCV repetitions. Categories are mutually exclusive and sum to 100% within each task. The sum of “All three incorrect” and “Three models correct” represents agreements; the sum of “One model correct” and “Two models correct” represents disagreements.

| **Cohort** | **Task** | **All three incorrect** | **One model correct** | **Two models correct** | **Three models correct** |
|---|---|---|---|---|---|
| **IRAP** | **APOE4** | 13.1% | 29.3% | 35.5% | 22.1% |
|  | **SUVR Q1** | 13.3% | 16.9% | 26.0% | 43.8% |
|  | **SUVR Median** | 18.8% | 28.2% | 31.1% | 21.9% |
|  | **SUVR Q4** | 15.1% | 22.0% | 28.9% | 34.0% |
| **WRAP** | **APOE4** | 20.7% | 18.7% | 23.6% | 37.0% |
|  | **PiB Q1** | 18.6% | 13.6% | 19.0% | 48.8% |
|  | **PiB Median** | 16.8% | 26.9% | 30.2% | 26.1% |
|  | **PiB Q4** | 13.5% | 9.2% | 17.0% | 60.3% |

Across all tasks, full agreement more often reflected cases in which all three models were correct than cases in which all three were incorrect. However, there was a substantial rate of disagreement, from 42.9% (SUVR Q1) to 64.8% (APOE4) in IRAP, and from 26.2% (PiB Q4) to 57.1% (PiB Median) in WRAP.

To examine whether correct predictions within disagreements were primarily attributable to a particular model or pair of models, we examined the six possible configurations for disagreements: only the cognitive, embedding, or top-down model correct, and only the cognitive–embedding, cognitive–top-down, or embedding–top-down pair correct. The contributions of these configurations were normalized by the total number of disagreement cases within each task (Figure 1). Across tasks and cohorts, no single constituent model or pair consistently accounted for the largest share of disagreements. Each configuration typically contributed approximately 11-23% of disagreement cases, with the greatest variability observed in WRAP PiB Q4, ranging from 7.6% (only top-down correct) to 26.3% (cognitive and top-down correct). The relatively even distribution across configurations suggests that disagreement patterns were not driven by one consistently superior constituent model or pair.

**Figure 1:**
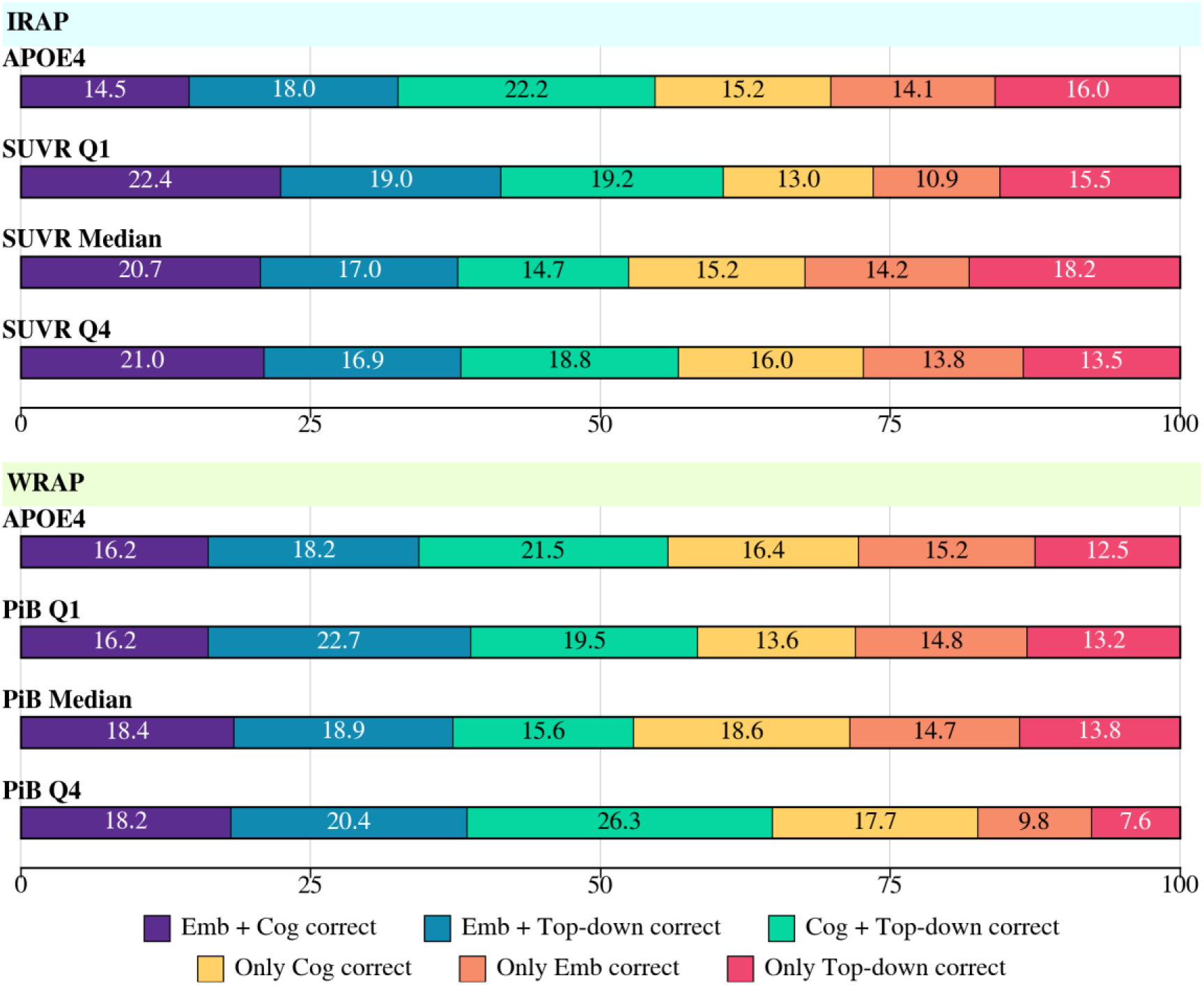
Constituent Model Contributions to Correct Predictions in Disagreements.

Class-specific analyses (Supplementary Table 7 and Supplementary Figure 1) showed broadly similar disagreement rates for positive and negative observations, with the largest difference, 17.0 percentage points, observed for IRAP SUVR Q1 (55.7% vs. 38.7% for positive and negative observations, respectively). However, the composition of disagreements differed: among positive observations, only one constituent model was typically correct in all tasks except IRAP SUVR Median, whereas among negative observations, two models were more often correct. No single constituent model consistently dominated the uniquely correct predictions among positive observations.

These findings demonstrate that the similar F1 scores achieved by the constituent models do not arise from correctly classifying the same observations. Rather, the models frequently succeed and fail on different observations, suggesting that they capture complementary predictive information.

Stacked bars show, for each prediction task in IRAP (top) and WRAP (bottom), how disagreement cases (test-set observations, pooled over 1,000 MCCV repetitions, for which only one or two constituent models were correct) were distributed across correct-prediction patterns.

Within each task, percentages were normalized by the total number of disagreements and sum to 100%. Colors distinguish the six possible patterns of correct predictions: three in which two constituent models were correct and three in which only one model was correct.

### 2.4 The Any-Positive Ensemble Outperforms Each Constituent Model

To assess whether the observed differences in individual-level predictions could be leveraged to improve classification performance, we implemented an any-positive ensemble that classified an observation as positive whenever at least one constituent model predicted a positive outcome. Across all four evaluated approaches, the any-positive ensemble consistently yielded the highest F1 score across all eight APOE4 and global amyloid tasks, with F1 scores of 0.435 (IRAP) and 0.427 (WRAP) for APOE4 and 0.345-0.640 (IRAP) and 0.352-0.573 (WRAP) for global amyloid, significantly outperforming every constituent model and the prior-informed random baseline, which assigned class labels according to their proportions in the task’s full sample (Table 4). In WRAP, the ensemble’s advantage was also observed when predicting clinically determined amyloid-positive versus amyloid-negative status, rather than outcomes defined using quartile-based thresholds (Supplementary Table 8).

**Table 4:** F1 Scores of Random Baseline, Constituent Models, and Any-Positive Ensemble Mean F1 scores across 1,000 MCCV repetitions, with 95% confidence intervals. Bold indicates the best-performing approach within each row. Asterisks denote statistically significant differences from the best-performing approach for the corresponding task, based on two-sided paired t-tests with BH FDR correction applied to the four comparisons within each task (* q<0.05, ** q<0.01, *** q<0.001). Test statistics and exact q values are reported in Supplementary Table 9.

| **Cohort** | **Task** | **Random (prior) [95% CI]** | **Cog [95% CI]** | **Emb [95% CI]** | **Top-down [95% CI]** | **Any positive [95% CI]** |
|---|---|---|---|---|---|---|
| **IRAP** | **APOE4** | 0.313<sup>***</sup><br>[0.30, 0.32] | 0.274<sup>***</sup><br>[0.26, 0.28] | 0.337<sup>***</sup><br>[0.33, 0.35] | 0.338<sup>***</sup><br>[0.33, 0.35] | **0.435<br>[0.43, 0.44]** |
|  | **SUVR Q1<br>(≤0.910)** | 0.236<sup>***</sup><br>[0.22, 0.25] | 0.334<sup>***</sup><br>[0.32, 0.35] | 0.285<sup>***</sup><br>[0.27, 0.30] | 0.294<sup>***</sup><br>[0.28, 0.31] | **0.417<br>[0.41, 0.43]** |
|  | **SUVR Median<br>(>0.941)** | 0.485<sup>***</sup><br>[0.47, 0.50] | 0.513<sup>***</sup><br>[0.50, 0.52] | 0.536<sup>***</sup><br>[0.53, 0.55] | 0.494<sup>***</sup><br>[0.48, 0.50] | **0.640<br>[0.63, 0.65]** |
|  | **SUVR Q4<br>(>0.980)** | 0.237<sup>***</sup><br>[0.22, 0.25] | 0.205<sup>***</sup><br>[0.19, 0.22] | 0.259<sup>***</sup><br>[0.25, 0.27] | 0.207<sup>***</sup><br>[0.19, 0.22] | **0.345<br>[0.33, 0.35]** |
| **WRAP** | **APOE4** | 0.368<sup>***</sup><br>[0.37, 0.37] | 0.260<sup>***</sup><br>[0.26, 0.26] | 0.294<sup>***</sup><br>[0.29, 0.30] | 0.263<sup>***</sup><br>[0.26, 0.27] | **0.427<br>[0.42, 0.43]** |
|  | **PiB Q1<br>(≤1.046)** | 0.294<sup>***</sup><br>[0.29, 0.30] | 0.180<sup>***</sup><br>[0.18, 0.18] | 0.281<sup>***</sup><br>[0.28, 0.28] | 0.260<sup>***</sup><br>[0.26, 0.26] | **0.372<br>[0.37, 0.38]** |
|  | **PiB Median<br>(>1.092)** | 0.436<sup>***</sup><br>[0.43, 0.44] | 0.460<sup>***</sup><br>[0.46, 0.46] | 0.475<sup>***</sup><br>[0.47, 0.48] | 0.450<sup>***</sup><br>[0.45, 0.45] | **0.573<br>[0.57, 0.57]** |
|  | **PiB Q4<br>(>1.236)** | 0.214<sup>***</sup><br>[0.21, 0.22] | 0.290<sup>***</sup><br>[0.29, 0.29] | 0.214<sup>***</sup><br>[0.21, 0.22] | 0.157<sup>***</sup><br>[0.15, 0.16] | **0.352<br>[0.35, 0.36]** |

### 2.5 The Any-Positive Ensemble Advantage Extends to Regional Amyloid Burden

We next examined whether the ensemble’s advantage extended beyond global amyloid measures to region-specific Aβ burden (Figure 2). We considered four cortical regions (cingulate, frontal, parietal, and temporal) and defined three binary tasks per region (Q1, above-median, and Q4 burden), using region-specific thresholds within each cohort. This yielded 12 regional classification tasks per cohort. In IRAP, these four cortical regions were those contributing to the global SUVR measure. In WRAP, the regional measures were derived from available anatomically grouped cortical regions and were not identical to the eight-region composite used for the global PiB Index (see Methods, Section 4.2.2).

**Figure 2:**
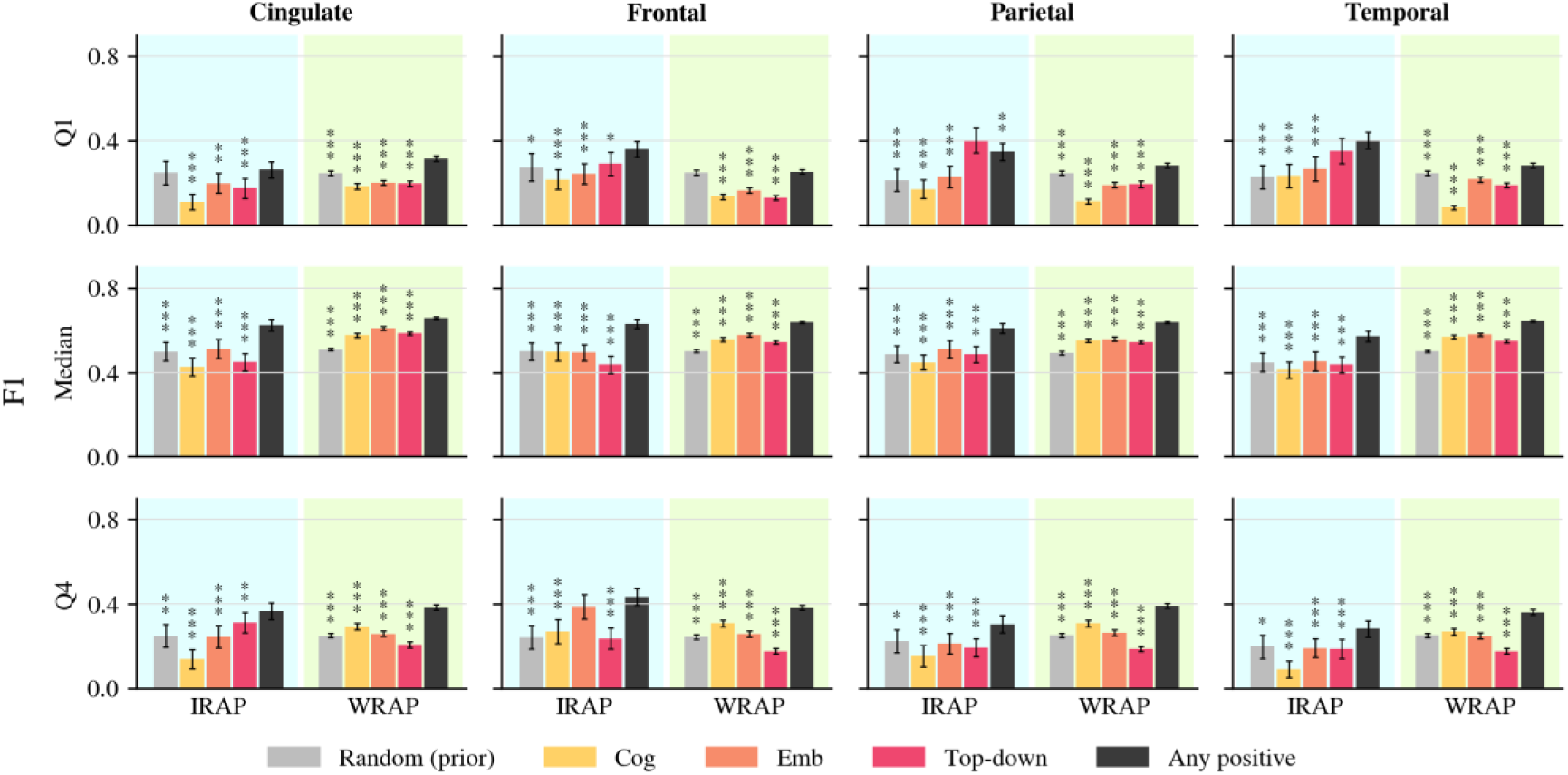
F1 Scores of Random Baseline, Constituent Models, and Any-Positive Ensemble for Regional Amyloid Prediction.

In WRAP, the any-positive ensemble achieved F1 scores ranging from 0.25 to 0.66 and significantly outperformed each of the three constituent models across all 12 regional tasks (two-sided BH-corrected paired t-tests, all q < 0.001). In IRAP, the ensemble achieved F1 scores ranging from 0.26 to 0.63 and significantly outperformed all three constituent models in 9 of the 12 tasks. Thus, the ensemble significantly outperformed every constituent model in 21 of the 24 regional tasks across both cohorts. In two of the remaining IRAP tasks, the ensemble had a nominally higher F1, but the difference from the strongest constituent model was not statistically significant (top-down model for temporal Q1, q = 0.066; embedding-based model for frontal Q4, q = 0.075). For parietal Q1, the top-down model significantly outperformed the ensemble (0.40 vs. 0.35, q = 0.008). In the tasks where the ensemble was the best-performing approach, it also significantly outperformed the prior-informed random baseline in all but two tasks (IRAP cingulate Q1, q = 0.680; WRAP frontal Q1, q = 0.622).

Mean F1 scores across 100 MCCV repetitions for the prior-informed random baseline, each constituent model, and the ensemble. Whiskers denote 95% confidence intervals. Asterisks denote statistically significant differences from the best-performing approach for the corresponding task, based on two-sided paired t-tests with BH FDR correction applied to the four comparisons within each task (* q<0.05, ** q<0.01, *** q<0.001). Test statistics and exact q values for each comparison are reported in Supplementary Table 10.

## 3 Discussion

This study examined whether language-based representations capture signals associated with preclinical AD risk and pathology, and whether these signals can enhance traditional cognitive assessments. We compared three sets of features: cognitive test scores, serving as the primary baseline for current clinical assessment; top-down, theory-driven linguistic features, representing the traditional approach to language analysis, and bottom-up linguistic embeddings derived from pretrained language models. Three main findings emerged. First, substantial individual-level disagreement among the constituent models indicated that each representation captured information not consistently recovered by the others. Second, embedding-based representations generally performed best among the constituent models, although no single representation consistently outperformed the others across all tasks. Third, combining the representations through an any-positive ensemble produced the best-performing model, significantly outperforming each constituent model and a prior-informed random baseline, and consistently yielding the highest F1 scores across both cohorts for APOE4 carrier status and global amyloid burden. This advantage also extended to regional amyloid burden, with the ensemble significantly outperforming all three constituent models in 21 of the 24 regional prediction tasks across four brain regions. Together, these findings suggest that linguistic representations and traditional cognitive measures capture complementary information associated with AD-related genetic risk and amyloid pathology, and that their integration improves predictive performance beyond that of individual representations.

This study provides a proof of concept for the potential utility of computational language analysis in preclinical AD research, with several implications for future clinical applications. First, because the Cookie Theft picture-description task is already incorporated into many cognitive assessments, our approach could extract additional information from an existing assessment with minimal additional effort. Second, language analysis could offer an accessible, low-burden screening approach to help identify individuals who may benefit from further clinical evaluation. Third, repeated speech collection could enable more frequent monitoring of linguistic changes, potentially even in home-based settings. This possibility is particularly relevant with the emergence of anti-amyloid treatments and the growing need for accessible methods of longitudinal monitoring. Although the present study did not examine within-person changes or treatment response, it provides a foundation for investigating whether language-based measures could serve as indicators of disease progression or response to treatment. Importantly, the clinical use of such automated tools should be accompanied by healthcare professionals who can explain their clinical meaning and uncertainty, provide appropriate support, and guide individuals toward confirmatory assessment or available preventive and therapeutic options when indicated. Language-based tools may therefore be most valuable as decision-support or screening instruments rather than as systems that communicate biological risk independently of clinical context.

This study shows consistent findings across two independent cohorts with substantially different linguistic and sociocultural characteristics. This consistency is particularly meaningful given the predominance of English-speaking populations in research on speech and language markers of neurodegeneration^31^, alongside evidence that linguistic alterations associated with AD and MCI may vary across languages^17,32–35^. By including Hebrew, an underrepresented language in this field, our study contributes to broadening the linguistic diversity of AD research and supporting the development of more linguistically inclusive and equitable screening tools. Although differences in data collection and biomarker measurement complicate direct comparisons, the convergence of our central findings despite these provide strong evidence for their generalizability across diverse populations and settings.

Our findings add to growing evidence linking language characteristics to Aβ pathology in cognitively unimpaired and mildly impaired individuals^14,36–39^, including previous work in the WRAP cohort^37^. Previous studies have also employed pretrained language models to predict amyloid positivity, either in samples including individuals with clinically manifest cognitive impairment^40,41^, and among cognitively unimpaired individuals^39^. However, these studies have framed amyloid prediction as binary classification based on predefined positivity criteria. Other studies have examined associations between language Aβ burden without relying on a binary clinical threshold, but have primarily focused on predefined linguistic or acoustic measures and did not employ LLM approaches^38,42^. Therefore, a key contribution of our study is the evaluation of amyloid burden across multiple distribution-based thresholds using LLM representations, rather than relying exclusively on a single clinical cutoff. This approach is particularly relevant given growing evidence that Aβ accumulation below conventional positivity thresholds may precede more rapid biomarker progression and subtle cognitive changes^43–45^. Our findings extend this evidence by suggesting that language-based representations capture signals associated with amyloid burden even below conventional thresholds for amyloid positivity.

Notably, disagreement in amyloid prediction was highest for the median-based classification tasks in both cohorts. This pattern is consistent with the possibility that more pronounced amyloid-related patterns are captured more similarly across the three approaches, whereas subtler signals lead to more divergent predictions. However, because median-based classification includes the full range of amyloid values and differs from quartile-based tasks in class balance, these findings do not establish that disagreement is concentrated specifically among individuals with intermediate amyloid burden. Future analyses examining disagreement as a function of continuous amyloid burden could help clarify these patterns.

This work also contributes to the relatively limited literature on spontaneous speech and APOE4 carrier status. Previous studies have often treated APOE4 as a covariate rather than a direct prediction target, including investigations of cognitive decline and AD-related biomarkers incorporating language or speech measures^12,37,42,46^. APOE4-related linguistic differences have also been examined using structured verbal neuropsychological tasks^46–48^. In contrast, attempts to infer APOE4 carrier status directly from spontaneous speech remain scarce. A recent study classified cognitively healthy APOE4 carriers and non-carriers using Cookie Theft recordings, but relied exclusively on acoustic characteristics^49^. By contrast, our study examines embedding-based and top-down linguistic representations, capturing multiple dimensions of language, including lexical, syntactic, and content-related characteristics.

Beyond the range of linguistic dimensions represented, another strength of this study lies in the quality of the underlying transcripts and annotations. Transcripts were carefully prepared by trained annotators following rigorous protocols. The IRAP transcripts were produced and meticulously annotated specifically for the present research, rather than relying on pre-existing transcriptions or automated transcription alone. This substantial data preparation effort involved independent annotation by two trained annotators, followed by adjudication of disagreements by a third annotator. Unlike approaches relying on readily available transcripts or uncorrected Automatic Speech Recognition (ASR) outputs, this procedure allowed careful preservation of subtle linguistic phenomena that may be particularly informative in preclinical AD research. The same emphasis on quality extended to the extraction of top-down linguistic features, which combined manual, automated, and LLM-based approaches. Disfluencies were manually annotated, an approach particularly valuable in clinical contexts, where subtle disruptions in speech may carry important signals of cognitive changes that off-the-shelf ASR systems may fail to capture reliably. Lexical measures were computed using language-specific NLP tools and corpus-based frequency norms, while higher-level, theory-driven features were annotated using a validated LLM-as-a-judge framework. These procedures allowed us to preserve linguistic phenomena that could otherwise be lost during preprocessing and to examine their potential relevance to AD-related biological measures.

Importantly, the predictive linguistic signals identified in this study were obtained using relatively simple and computationally accessible approaches. LLM embeddings were extracted from pretrained, off-the-shelf language models with frozen weights, without additional fine-tuning. The top-down approach relied on only 13 theory-driven linguistic features, selected to represent diverse dimensions of language, including lexical, syntactic, and content-related characteristics. Both representations were used as inputs to HMDR, a linear classifier. Similarly, our ensemble employed a simple decision rule, classifying an observation as positive if at least one constituent model predicted a positive outcome, without additional training or parameter optimization. The ability to combine lightweight approaches to predict AD-related indicators has important implications for future clinical applications, as their modest computational demands not only safeguard patient privacy through local deployment, but also ensure accessibility and reproducibility in resource-limited settings.

These results establish a foundation for further investigating the predictive value of spontaneous speech in preclinical AD, while demonstrating that meaningful signals can be extracted using relatively simple linguistic representations. At the same time, the current approaches should not be considered an upper bound on achievable performance. Future work could examine whether larger or task-adapted language models, fine-tuning with sufficiently large datasets, or broader sets of linguistically informed features provide additional predictive value. Such developments should be evaluated against the interpretability, efficiency, and accessibility of the relatively lightweight approaches demonstrated here.

Several limitations of this study should be acknowledged. First, although WRAP includes repeated speech observations for some participants, the present analyses do not model within-person linguistic change over time. Accordingly, the threshold-specific patterns cannot be interpreted as longitudinal trajectories of language change as Aβ accumulates. Future repeated-measures analyses linking within-person changes in language and biomarkers are needed to test this directly.

Second, methodological and cohort-specific differences between IRAP and WRAP limit our ability to separate the observed associations between linguistic and cognitive measures and the target variables from potential influences of differences in data collection, biomarker measurement, and social and cultural characteristics. IRAP and WRAP are active longitudinal cohorts whose protocols continue to evolve, creating differences in available measures and participant overlap across modalities. We therefore focused on APOE4 and Aβ, the two AD-related biological indicators currently available in both cohorts with sufficient overlap with Cookie Theft speech samples. Other potentially informative markers are either available in only one cohort or do not yet have sufficient overlap with the speech data. Particularly relevant future targets include tau and blood-based biomarkers such as GFAP and NfL, which are being collected as these cohorts continue to expand.

Although cognitive inputs were initially defined according to the established literature for each cohort, an additional evaluation using a harmonized set of cognitive measures available in both cohorts showed that the pattern remained largely unchanged, with no consistently superior constituent model and the ensemble outperforming all constituent models across tasks (Supplementary Methods 1.2 and Supplementary Tables 3-5). Nevertheless, other sources of cross-cohort variability remain, including differences in amyloid tracers and processing procedures, cultural and sociolinguistic factors that may shape language use, and potential differences in how embedding models represent Hebrew and English. Harmonizing amyloid measures using Centiloid scaling, together with validation across additional languages and cohorts, could help disentangle these sources of variability. Importantly, the consistency of these findings across two independent cohorts, despite linguistic, sociocultural, and methodological differences, further strengthens the evidence for the complementarity of linguistic and cognitive measures and the predictive benefits of integrated approaches.

## 4 Methods

### 4.1 Participants

Participants were drawn from two longitudinal cohorts focused on preclinical AD, IRAP and WRAP. The cohorts comprised, respectively, middle-aged Hebrew- and English-speakers with a parental history of AD. Available data included speech recordings, cognitive assessments, neuroimaging, genetic information, and electronic health records. IRAP data were consolidated from several separate data files spanning June to September 2025; WRAP data were obtained from the November 30, 2025 data freeze. All participants provided written informed consent. Data collection was approved by the Institutional Review Board of Sheba Medical Center for IRAP and by the University of Wisconsin-Madison Institutional Review Board for WRAP.

To compare the language and cognitive models, we included observations for which both language and cognitive data were available, together with APOE4 carrier status, Aβ burden, or both. The cognitive assessment and Cookie Theft task were typically completed on the same day. However, these assessments could be separated in time from the Positron Emission Tomography (PET) scan used to quantify amyloid burden. In both cohorts, PET scans were restricted to within ±8 years of the assessment, limiting the temporal gap while allowing to retain all available IRAP observations.

In both IRAP and WRAP cohorts, participants contributed speech samples across multiple visits. For amyloid analyses, each speech sample was paired with the temporally closest eligible PET/PiB scan, allowing individual scans to serve multiple observations (IRAP: 59 observations across 58 unique scans; WRAP: 1,182 observations across 559 unique scans). Because APOE4 genotype is time-invariant, single genotype calls were mapped across all repeated visits for each participant (IRAP: 126 observations from 125 unique calls; WRAP: 1,538 observations from 538 unique calls).

In IRAP, participants were classified using the Clinical Dementia Rating (CDR)^50^ into Healthy Control (HC; n = 120) and MCI (n = 14) groups. In WRAP, cognitive status was determined through a two-tiered review process, in which visit data were initially screened by an algorithm, followed by multidisciplinary consensus review of flagged cases^51,52^. This process resulted in one of five classifications: Cognitively Unimpaired-Stable (CU-S; n=1,265), Cognitively Unimpaired-Declining (CU-D; n=222), MCI (n=46), impaired but not MCI (n=5), and dementia (n=3). CU-S denotes individuals with normal cognition and no evidence of meaningful decline, whereas CU-D denotes individuals with evidence of cognitive decline that is not sufficient to meet criteria for MCI. The impaired-but-not-MCI category includes individuals with cognitive impairment that was not classified as either CU-D or MCI, including cases in which impairment was considered longstanding rather than a decline from a previous level of functioning. For cross-cohort comparison, cognitive status was harmonized into two categories: cognitively unimpaired (IRAP: HC; WRAP: CU-S and CU-D) and cognitively impaired (IRAP: MCI; WRAP: MCI, impaired but not MCI, and dementia).

### 4.2 Target variables

#### 4.2.1 APOE4

APOE4 carrier status was treated as a binary classification task, defined as the presence of at least one ε4 allele of the APOE gene. In both cohorts, APOE genotype information was obtained from blood samples.

#### 4.2.2 Amyloid-β

Both cohorts used PET combined with Computed Tomography (PET-CT) scans to quantify amyloid tracer accumulation: the SUVR in IRAP, and PiB Index in WRAP. Further details are provided later in this section.

To examine whether language and cognitive measures captured signals across different levels of amyloid accumulation, three binary classification tasks were defined separately for each cohort using cohort-specific thresholds: membership in the lowest quartile, values above the median, and membership in the highest quartile. These tasks were defined for the global amyloid measure in each cohort and, separately, for each of four regional amyloid measures (cingulate, frontal, parietal, and temporal). For the regional tasks, the thresholds were determined independently from the distribution of the corresponding region within each cohort, rather than reusing the global biomarker cutoffs. Each cohort therefore contributed three global amyloid tasks and 12 regional amyloid tasks.

Clinical practice routinely relies on expert interpretation to determine the presence or absence of amyloid pathology. Therefore, we additionally evaluated the prediction of this binary status in WRAP (Supplementary Table 8). IRAP was excluded from this analysis due to severe class imbalance, with only two positive cases.

##### 4.2.2.1 IRAP

As previously described^28^, SUVR values were derived using PET-CT. SUVR reflected concentration of the ^18^F tracer in cortical target regions (cingulate gyrus, frontal, parietal, temporal cortices) relative to the cerebellum, a reference region typically spared from amyloid deposition even in advanced stages of AD. Global SUVR was calculated as a weighted average of the SUVR values across these four cortical regions. The three binary classification tasks were defined using the global IRAP SUVR distribution: membership in the first quartile (SUVR < 0.910), the upper median (SUVR > 0.941), and the fourth quartile (SUVR > 0.980).

For the regional analyses, the four cortical SUVR values (cingulate, frontal, parietal, and temporal) were used as separate outcomes. The Q1, median, and Q4 thresholds were calculated independently for each regional SUVR distribution, rather than using the global SUVR thresholds, resulting in the following thresholds: cingulate, Q1 < 0.998, median > 1.043, Q4 > 1.089; frontal, Q1 < 0.907, median > 0.938, Q4 > 0.982; parietal, Q1 < 0.920, median > 0.945, Q4 > 0.995; temporal, Q1 < 0.880, median > 0.923, Q4 > 0.949. Due to differences in data availability, the regional analyses included fewer scans than the global SUVR analysis (N = 59), with 41 observations available for the cingulate, parietal, and temporal regions and 40 for the frontal region.

##### 4.2.2.2 WRAP

Amyloid burden was quantified via 70-minute [¹¹C]PiB PET scans. The dynamic PET data were transformed into voxel-wise distribution volume ratio (DVR) maps using the cerebellar cortex as the reference region. Cortical DVR values from eight regions-anterior and posterior cingulate, medial orbitofrontal, supramarginal, angular, middle temporal, superior temporal, and precuneus-were then averaged into the composite PiB Index per the methodology detailed in ^53–55^. The three binary classification tasks were then defined from the distribution of PiB Index values in WRAP: membership in the first quartile (PiB Index < 1.046), values above the median (PiB Index > 1.092), and membership in the fourth quartile (PiB Index > 1.236).

In WRAP, regional DVR values were extracted using the Automated Anatomical Labeling (AAL) atlas and grouped into the four broad cortical regions examined in IRAP: frontal (superior and middle frontal gyri; opercular and triangular parts of the inferior frontal gyrus; medial superior frontal gyrus; orbital portions of the superior, middle, and inferior frontal gyri; medial orbitofrontal cortex; and gyrus rectus), parietal (superior and inferior parietal lobules, angular and supramarginal gyri, and precuneus), temporal (superior, middle, and inferior temporal gyri), and cingulate (anterior, middle, and posterior cingulate gyri). These regional measures incorporated additional cortical DVR values beyond the eight regions used to calculate the global PiB Index. The Q1, median, and Q4 thresholds were calculated independently for each regional DVR distribution, resulting in the following thresholds: cingulate, Q1 < 1.124, median > 1.176, Q4 > 1.319; frontal, Q1 < 1.002, median > 1.041, Q4 > 1.161; parietal, Q1 < 0.981, median > 1.021, Q4 > 1.147; and temporal, Q1 < 1.004, median > 1.038, Q4 > 1.123.

### 4.3 Experimental setup

We compared three constituent prediction models-cognitive, embedding-based, and top-down-each combining a distinct feature representation with the same HMDR framework. An analysis of agreement and disagreement among their predictions motivated evaluation of an “any-positive” ensemble strategy, which classified an observation as positive if at least one constituent model predicted a positive outcome. The model comparison was carried out for APOE4 and global amyloid outcomes and was extended to the four regional amyloid measures, using the cohort-specific classification thresholds defined above.

The following subsections describe the speech task and preprocessing procedure (4.3.1), the three constituent model input representations (4.3.2), constituent model-development procedure (4.3.3), and performance evaluation (4.3.4).

#### 4.3.1 Speech Task and Preprocessing

Participants completed the Boston Cookie Theft picture-description task^8^, a narrative elicitation task widely used in cognitive assessment. IRAP participants were shown the picture presented in Supplementary Figure 2 and instructed to “describe the picture with as many details as possible” (translated from Hebrew). WRAP participants completed the same picture-description task with the instruction “Tell me everything you see going on in this picture”.

IRAP recordings were transcribed using Subtitle Edit 4.0.11 by two blinded annotators, achieving substantial inter-annotator agreement (Cohen’s κ = 0.758). Discrepancies were resolved by a third adjudicator.

WRAP transcripts were annotated in CHAT format by trained speech-language pathology graduate students from the Mueller Lab at the Center for Communication and Neurodegenerative Disease and Language (CCANDL), University of Wisconsin-Madison, as described in ^13^.

Interviewer speech, and non-verbal vocalizations (e.g., laughter, coughing) and unintelligible words were removed. Filler words, repetitions, and false starts were retained to preserve spontaneous speech characteristics.

#### 4.3.2 Constituent Model Inputs

##### 4.3.2.1 Cognitive Model

The cognitive model was based on neuropsychological test batteries administered in the IRAP and WRAP cohorts. Raw test scores were standardized to z-scores prior to modeling. Previous studies within each cohort have also defined composite scores that aggregate individual tests into cognitive domains, as well as global measures of cognitive performance. Accordingly, we evaluated cognitive features at three levels of representation: individual standardized test scores, domain-level composite scores, and global cognitive scores. The specific tests and composite definitions included at each level are detailed in Supplementary Methods 1.1.

##### 4.3.2.2 Embedding Model

For each transcript, embeddings were extracted independently from each language model included for the corresponding cohort, yielding multiple candidate representations of the same text. These representations were treated as a hyperparameter and selected within the model-training procedure, detailed in Supplementary Methods 3.2. All models were downloaded and executed locally to protect participant privacy and data security. The embedding models spanned multilingual, English-specific, and Hebrew-specific architectures and varied in pretraining objectives, model size, and embedding dimensionality, as described in Supplementary Methods 2.1. By incorporating both multilingual and language-specific architectures, this framework facilitates direct cross-cohort comparisons between English and Hebrew while maximizing representation quality within the Hebrew-speaking cohort.

##### 4.3.2.3 Top-Down Model

In contrast to embedding-based representations, which encode text through latent semantic patterns that are not directly interpretable, the top-down approach relies on predefined, theory-driven features with explicit meaning, grounded in the clinical and linguistic literature. Table 5 outlines this feature set, including extraction methods for the IRAP and WRAP cohorts, across five categories: disfluencies, lexical and syntactic properties, objective content, subjective interpretation, and human experience.

**Table 5:** Features Included in the Top-Down Analysis Features included in the top-down analysis, grouped by category, with the corresponding extraction method for the IRAP and WRAP cohorts.

| **Category** | **Feature** | **IRAP Extraction Method** | **WRAP Extraction Method** |
|---|---|---|---|
| Disfluencies | # Fillers | Manual annotation | Manual annotation |
|  | # False starts | Manual annotation | Manual annotation |
|  | # Repetitions | Manual annotation | Manual annotation |
| Lexical and Syntactic Properties | Open/Closed class ratio | Automatic count based on DictaBERT<sup>56</sup> POS tagging | Automatic count based on CLAN pipelines |
|  | Pronoun/Pronoun+Noun ratio | Automatic count based on DictaBERT<sup>56</sup> POS tagging | Automatic count based on CLAN pipelines |
|  | Type-Token Ratio (TTR) | Automatic count based on DictaBERT<sup>56</sup> POS tagging | Automatic count based on CLAN pipelines |
|  | Frequency | wordfreq 3.1.1<sup>59</sup> Hebrew norms | wordfreq 3.1.1<sup>59</sup> English norms |
|  | Short Sentences | LLM | LLM |
| Objective Content | Correct Information Units (CIU) | LLM | Manual annotation |
| Subjective Interpretation | Theory of Mind Mother | LLM | LLM |
|  | Theory of Mind Boy | LLM | LLM |
|  | Theory of Mind Girl | LLM | LLM |
| Human Experience | Criticize Characters | LLM | LLM |

*Disfluencies* captured disruptions in language production and were derived from manual transcript annotation followed by automated counting. *Lexical and syntactic properties* captured word- and sentence-level characteristics and were extracted primarily using conventional computational methods, including automatic linguistic analysis and language-specific lexical resources: For IRAP, tokenization and Part-of-Speech (POS) tagging were performed using DictaBERT^56^, an open-source NLP toolkit that provides linguistic analyses including tokenization, POS tagging, and dependency parsing. For WRAP, corresponding features were derived using CLAN (Computerized Language ANalysis)^57,58^ pipelines under the transcription and analysis conventions maintained by TalkBank. Word-frequency in both Hebrew and English was derived using the Python wordfreq library (version 3.1.1)^59^, which provides corpus-based estimates of word frequency across multiple languages.

In addition, using a previously developed LLM-based annotation framework^60,61^, we extracted a set of high-level binary features defined across four categories: *lexical and syntactic properties, objective content, subjective interpretation,* and *human experience*. These features assess the description as a whole rather than isolated words, sentences, or directly computed counts. Developed in consultation with experts and grounded in previous dementia research, they characterize not only what information the speaker reports, but also how the speaker interprets and responds to the depicted situation. Therefore, this approach complements conventional quantitative language measures by capturing complex, interpretable behaviors that are difficult to operationalize through simple counts or rule-based algorithms.

To enable these high-level features to be annotated at scale, we used Qwen3.5-9B^62^ as the LLM annotator. The quality of the LLM annotations was evaluated using the Alternative Annotator Test (AAT)^63^, a statistical method for assessing whether annotations produced by an LLM can serve as a substitute for human annotations (Supplementary Methods 2.4 and Supplementary Table 11). WRAP Correct Information Units (CIU) were already available from manual annotation, obviating the need for LLM annotation.

Detailed feature definitions and annotation prompts are provided in Supplementary Methods 2.2. Additional preliminary analyses, including descriptive statistics and exclusion criteria for features are provided in Supplementary Methods 2.3, Supplementary Table 12, and Supplementary Figure 3.

#### 4.3.3 Model Development

Model development for all classification tasks was conducted using stratified MCCV and HMDR, which captures shared population trends and domain-specific variations. The HMDR formulation is detailed in Supplementary Methods 3.1. In each iteration, 20% of the samples were randomly held out as an independent test set, while the remaining 80% were used for hyperparameter tuning. All observations belonging to the same participant were assigned to the same partition. In the APOE4 analyses, the feature vector contained either cognitive, embedding, or top-down features. In amyloid-related tasks, both in IRAP and WRAP, the interval in years between the speech or cognitive assessment and the matched PET-CT scan was appended to the feature vector. All features, including the scan-speech gap, were standardized within the training data before model fitting. Hyperparameter selection was performed exclusively within the training set in two sequential phases (Supplementary Methods 3.2), each using a separate stratified five-fold cross-validation procedure, with hyperparameters selected to maximize the positive-class F1 score.

#### 4.3.4 Evaluation

Using F1 as the primary evaluation metric, we compare the constituent models with the any-positive ensemble to assess whether combining multiple representations yields a significant improvement in predictive performance. As a reference for chance-level performance, all approaches were also compared with a prior-informed random baseline, which assigned class labels according to their proportions in the task’s full sample. The F1 score is defined as:

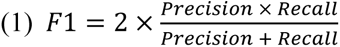

Where:

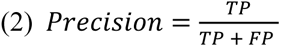

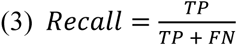

TP, FP, and FN denote true positives, false positives, and false negatives, respectively.

Precision, also known as Positive Predictive Value (PPV), measures how many of all cases predicted as positive are truly positive. Recall, also known as sensitivity or True Positive Rate (TPR), measures how many of all actual positive cases are correctly identified by the model.

The F1 score combines precision and recall into a single measure, thereby accounting for both false-positive and false-negative errors. By definition, it is their harmonic mean, therefore a low value for either precision or recall substantially reduces the overall F1 score. Additional performance metrics, including precision, recall, area under the receiver operating characteristic curve (ROC-AUC), and area under the precision-recall curve (PR-AUC), are reported in Supplementary Methods 3.3 and Supplementary Tables 13-16.

### 4.4 Statistical analysis

Continuous demographic and clinical characteristics were summarized as mean (SD), and categorical variables as counts and percentages. Differences between cohorts and between positive and negative outcome groups within each classification task were assessed using two-sided Welch’s t-tests for continuous variables and Pearson’s chi-square tests for categorical variables.

Predictive performance was evaluated across 1,000 MCCV repetitions for APOE4 and global Aβ classification tasks, and 100 repetitions for regional Aβ tasks. F1 scores were calculated separately for each outcome class on the held-out test sets and summarized as the mean across repetitions. Mean F1 scores were reported with 95% confidence intervals. Additional performance metrics included precision, recall, ROC-AUC, and PR-AUC (Supplementary Methods 3.3).

Pairwise comparisons of model performance were conducted using F1 scores obtained from matched MCCV test partitions. For *APOE4* and global Aβ tasks, comparisons proceeded in two stages. First, the highest-performing constituent model was evaluated against the remaining two constituent models. Second, the top-performing overall approach (consistently the “any-positive” ensemble in both tasks) was compared against the other four modeling approaches (the three constituent models and the prior-informed random baseline). For regional Aβ tasks, only this second stage was conducted, comparing the best-performing approach with the remaining four approaches among the three constituent models, the ensemble, and the prior-informed random baseline. Differences were assessed using two-sided paired t-tests, with BH FDR correction applied separately to the comparisons within each task and stage (two comparisons for constituent models, four for the full model comparison in APOE4, global Aβ, and regional Aβ tasks). Full test statistics are reported in Supplementary Tables 6, 9, and 10. Statistical significance was defined as q < 0.05.

## Data Availability

All data produced in the present study are available upon reasonable request to the authors.

https://wrap.wisc.edu/data-requests-2/

## Data Availability

WRAP data can be requested through an online submission process via https://wrap.wisc.edu/data-requests-2/. IRAP data may be made available to researchers upon request to the study’s principal investigator (R.R.S.).

## Author Contribution

M.Z., N.C., Y.W., R.R.S., M.S.B. and R.R. contributed to the conceptualization and design of the study. G.A. and T.S. were responsible for IRAP speech recordings, while R.A., S.L., G.N., and G.L. conducted IRAP transcript annotations. M.Z. drafted the manuscript. Y.W., N.R., I.G., O.L.S., K.D.M., R.R.S., M.S.B., and R.R. provided feedback and revised the manuscript. K.D.M. provided expertise on the WRAP speech and language data and contributed to their interpretation. R.R.S. served as the principal investigator of IRAP. M.S.B. and R.R. supervised the project.

